# Discontinuation of palliative brain radiotherapy in patients with brain metastases

**DOI:** 10.1101/2023.10.21.23297351

**Authors:** Paul Windisch, Jamie Lütscher, Robert Förster, Daniel R. Zwahlen, Christina Schröder

## Abstract

Discontinuation of radiotherapy is rarely discussed in the literature. In this retrospective study of 468 consecutive patients receiving palliative radiotherapy for brain metastases, we identified 35 discontinued treatments, mainly due to clinical deterioration. Poor performance status, more advanced disease and, in turn, poor prognosis was associated with higher discontinuation rates.

## Introduction

Treatment discontinuation in radiation oncology is rarely discussed in the scientific literature. A potential reason could be that a common cause of treatment discontinuation is the clinical deterioration of the patient and that, therefore, presenting cases of treatment discontinuation from one’s own institution might be viewed as a failure to perform good patient selection. However, good patient selection is difficult, especially when treating brain metastases where the prognosis varies greatly for different primary tumors and patient characteristics [1]. In addition, the improval of CNS-active systemic therapies has contributed to improved survival in some but not all patients with sometimes exceptional responses in patients with initially poor performance status, further complicating the issue [2,3]. However, good patient selection is crucial, especially since not every patient will derive a meaningful benefit, even from completing a course of brain radiotherapy as illustrated by the QUARTZ trial that found only a marginal difference in overall survival and quality-adjusted life-years (QALYs) for whole brain radiotherapy plus optimal supportive care compared to optimal supportive care alone [4].

Increasing the amount of data on the topic could potentially allow for better patient selection in the future. We, therefore, conducted a retrospective review of patients who were treated with cranial radiotherapy for brain metastases from solid tumors at our department. Our hypothesis was that treatment discontinuations would occur more frequently in patients with worse prognosis and the goal was in turn to identify characteristics frequently associated with treatment discontinuation.

## Methods

We retrospectively reviewed all patients who were treated with cranial radiotherapy for brain metastases from solid tumors in our department from 01/2010 to 12/2020. Patients who only received prophylactic cranial radiotherapy, had primary brain tumors, or hematologic malignancies were excluded. If a patient did not complete the number of therapy sessions that were prescribed at the start of the treatment, this was considered a case of treatment discontinuation.

In addition to collecting relevant patient characteristics and collecting the reasons for treatment discontinuation, we also computed the Recursive Partitioning Analysis (RPA) and disease-specific Graded Prognostic Assessment (GPA) groups for each patient where all the required information was available [5,6].

Chi square tests were used to test the frequencies of treatment discontinuations across the different RPA and GPA groups.

Data preprocessing, analysis, and visualization were performed with Python (version 3.9.7) using the numpy (version 1.20.3), pandas (version 1.3.4), scikit-learn (version 0.24.2), scipy (version 1.10.0), matplotlib (version 3.4.3), and seaborn (version 0.11.2) packages. The full dataset, notebook and environment file have been uploaded to a public repository (https://github.com/windisch-paul/rt-treatment-discontinuation).

Institutional review board approval was obtained from the ethical review committee of the canton of Zurich for a project (project number: BASEC 2020-02112) to analyze the effects and side effects of radiotherapy at our institution (ClinicalTrials.gov Identifier: NCT05192876). Written informed consent for the analysis of anonymized clinical and imaging data was obtained from all patients, and all data were gathered in accordance with the World Medical Association Declaration of Helsinki: Research involving human subjects.

## Results

The patient characteristics are presented in table 1 and the distributions of selected patient characteristics are presented in Figure 1A. Out of 468 patients who underwent cranial radiotherapy, 35 treatments (7.5%) were discontinued. The most frequent reason for treatment discontinuation was clinical deterioration which was documented in 26 (74.3%) of patients. One patient died, one patient underwent surgery for his brain metastases, and one patient discontinued his radiotherapy in order to start another systemic therapy. In the six remaining patients, no definitive reason was documented.

**Table 1.**
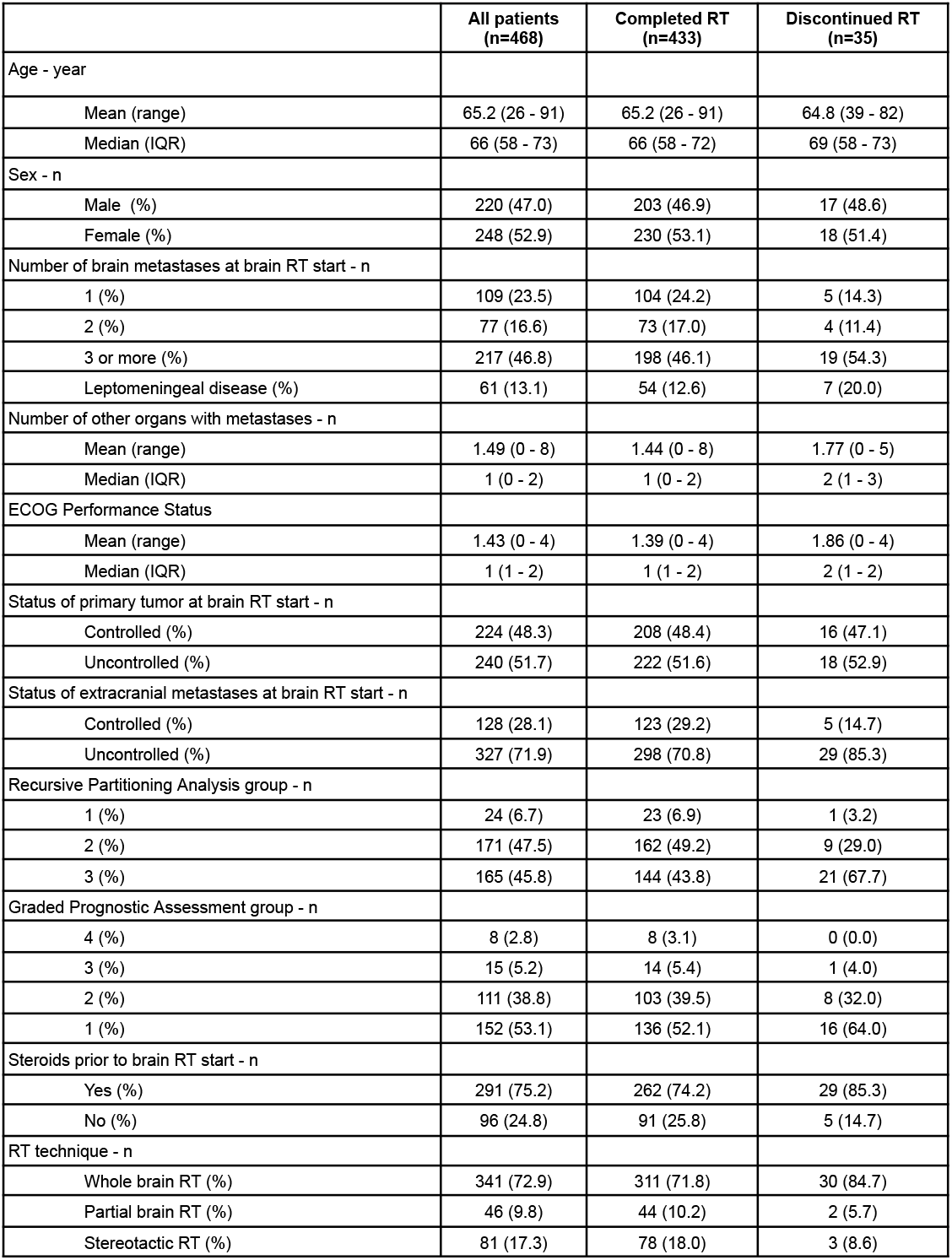
Patient characteristics. *RT = Radiotherapy, IQR = Interquartile range, ECOG = Eastern Cooperative Oncology Group*.

|  | All patients<br>(n=468) | Completed RT<br>(n=433) | Discontinued RT<br>(n=35) |
| --- | --- | --- | --- |
| Age - year |  |  |  |
| Mean (range) | 65.2 (26 - 91) | 65.2 (26 - 91) | 64.8 (39 - 82) |
| Median (IQR) | 66 (58 - 73) | 66 (58 - 72) | 69 (58 - 73) |
| Sex - n |  |  |  |
| Male (%) | 220 (47.0) | 203 (46.9) | 17 (48.6) |
| Female (%) | 248 (52.9) | 230 (53.1) | 18 (51.4) |
| Number of brain metastases at brain RT start - n |  |  |  |
| 1 (%) | 109 (23.5) | 104 (24.2) | 5 (14.3) |
| 2 (%) | 77 (16.6) | 73 (17.0) | 4 (11.4) |
| 3 or more (%) | 217 (46.8) | 198 (46.1) | 19 (54.3) |
| Leptomeningeal disease (%) | 61 (13.1) | 54 (12.6) | 7 (20.0) |
| Number of other organs with metastases - n |  |  |  |
| Mean (range) | 1.49 (0 - 8) | 1.44 (0 - 8) | 1.77 (0 - 5) |
| Median (IQR) | 1 (0 - 2) | 1 (0 - 2) | 2 (1 - 3) |
| ECOG Performance Status |  |  |  |
| Mean (range) | 1.43 (0 - 4) | 1.39 (0 - 4) | 1.86 (0 - 4) |
| Median (IQR) | 1 (1 - 2) | 1 (1 - 2) | 2 (1 - 2) |
| Status of primary tumor at brain RT start - n |  |  |  |
| Controlled (%) | 224 (48.3) | 208 (48.4) | 16 (47.1) |
| Uncontrolled (%) | 240 (51.7) | 222 (51.6) | 18 (52.9) |
| Status of extracranial metastases at brain RT start - n |  |  |  |
| Controlled (%) | 128 (28.1) | 123 (29.2) | 5 (14.7) |
| Uncontrolled (%) | 327 (71.9) | 298 (70.8) | 29 (85.3) |
| Recursive Partitioning Analysis group - n |  |  |  |
| 1 (%) | 24 (6.7) | 23 (6.9) | 1 (3.2) |
| 2 (%) | 171 (47.5) | 162 (49.2) | 9 (29.0) |
| 3 (%) | 165 (45.8) | 144 (43.8) | 21 (67.7) |
| Graded Prognostic Assessment group - n |  |  |  |
| 4 (%) | 8 (2.8) | 8 (3.1) | 0 (0.0) |
| 3 (%) | 15 (5.2) | 14 (5.4) | 1 (4.0) |
| 2 (%) | 111 (38.8) | 103 (39.5) | 8 (32.0) |
| 1 (%) | 152 (53.1) | 136 (52.1) | 16 (64.0) |
| Steroids prior to brain RT start - n |  |  |  |
| Yes (%) | 291 (75.2) | 262 (74.2) | 29 (85.3) |
| No (%) | 96 (24.8) | 91 (25.8) | 5 (14.7) |
| RT technique - n |  |  |  |
| Whole brain RT (%) | 341 (72.9) | 311 (71.8) | 30 (84.7) |
| Partial brain RT (%) | 46 (9.8) | 44 (10.2) | 2 (5.7) |
| Stereotactic RT (%) | 81 (17.3) | 78 (18.0) | 3 (8.6) |

**Figure 1.**
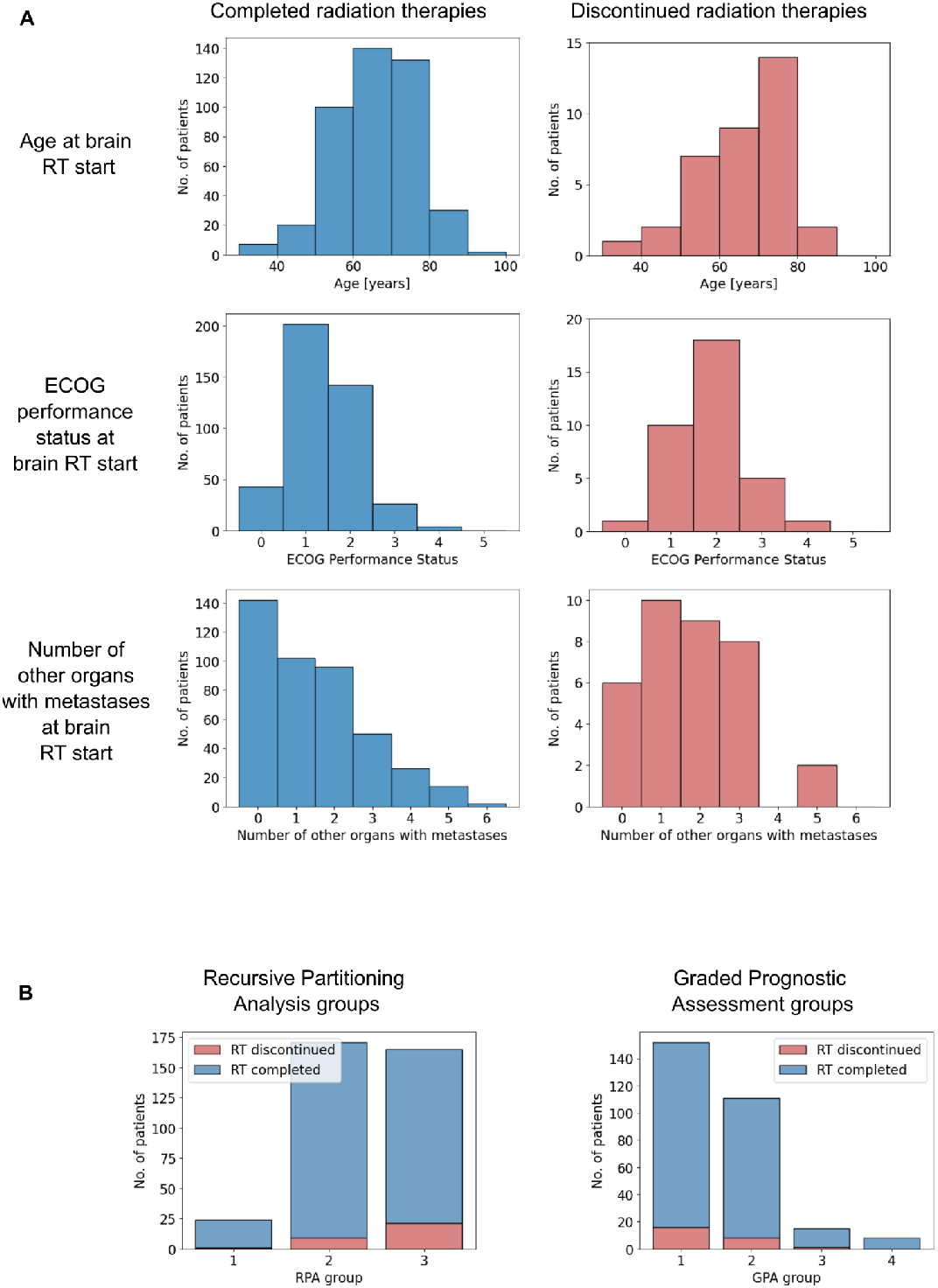
A) Histograms of selected patient characteristics depending on whether the radiotherapy was completed (left) or discontinued (right). B) Histograms of completed and discontinued radiotherapies by Recursive Partitioning analysis (left) and Graded Prognostic Assessment group (right). *RT = Radiotherapy, ECOG = Eastern Cooperative Oncology Group, RPA = Recursive Partitioning Analysis, GPA = Graded Prognostic Assessment*.

Patients whose radiotherapy was discontinued had, on average, more leptomeningeal disease (20.0% vs. 12.6%), worse ECOG performance status (mean ECOG performance status 1.86 vs. 1.39) and more uncontrolled extracranial metastases (85.3% vs. 70.8%). In addition, they were slightly older (median age 69 years vs. 66 years), more frequently diagnosed with three or more brain metastases (54.3% vs. 46.1%), more frequently under treatment with steroids (85.3% vs. 74.2%), and more frequently treated with whole brain radiotherapy (84.7% vs. 71.8%) instead of stereotactic or partial brain radiotherapy.

The distribution of RPA and GPA groups and the respective frequencies of treatment discontinuation are presented in figure 1B. The majority of patients were classified into the RPA groups 2 and 3 (47.5% and 45.8% respectively) which indicates an on average poor prognosis of the patient collective. For the GPA, this was even more pronounced with 64.0% of patients being classified in group 1 which is associated with the worst prognosis. The frequencies of treatment discontinuation increased with worse prognosis and differed significantly across RPA groups (p = 0.037) but not across GPA groups (p = 0.612).

## Discussion

Our analysis did not identify a singular factor that predicts the discontinuation of cranial radiotherapy for brain metastases from solid tumors. However, treatment discontinuations were more frequent in patients with generally poor performance status and more advanced or uncontrolled disease.

Due to the low number of treatment discontinuation events, we refrained from trying to build our own model to predict treatment discontinuation and instead attempted a validation of the performance of previously published prognostic models for the purpose of predicting treatment discontinuation.

As expected, we found an increase in rates of treatment discontinuation in the groups with worse prognosis for both, the GPA and the RPA. This also fits to clinical deterioration accounting for the majority of discontinued treatments. The fact that the frequency of treatment discontinuation across prognostic groups did not reach statistical significance for the GPA is likely due to the low number of discontinuation events combined with the higher number of classes compared to the RPA.

Searching PubMed for reports on radiotherapy discontinuation (query syntax: “((discontinuation[Title]) OR (stop[Title]) OR (termination[Title]) OR (abort[Title])) AND ((radiotherapy[Title]) OR (radiation[Title]))”) yielded 44 results, four of which were original articles discussing the permanent discontinuation of radiotherapy [7–10]. Lebwohl and colleagues analyzed treatment discontinuation during radiochemotherapy for rectal cancer and found a discontinuation rate of 5.3% [9]. Ramsey and colleagues found that 22% of Medicare-enrolled women did not complete their radiotherapy for non-metastatic breast cancer [10]. Puckett and colleagues analyzed 297 patients who received any kind of palliative radiotherapy and found that 60 (20.2%) did not complete their treatment and also report an association of worse performance status with increased rates of discontinuation. Unsurprisingly, they also saw a correlation of treatment discontinuation with poor survival [8]. Lazarev and colleagues reported that 58 out of 1,001 patients (5.7%) discontinued their curative radiotherapy for head and neck cancer due to a variety of different reasons, from patients’ decisions against medical advice (33%), over comorbidities (24%), toxicities (17%), social factors (17%) to disease progression (9%) [7]. The discrepancy between the reasons they found in a head and neck cohort compared to the reasons we identified in a brain metastases cohort suggests that reasons for treatment discontinuation and in turn factors predicting the risk of treatment discontinuation might vary greatly for different tumor sites and clinical scenarios.

The strengths of our article include the consecutive sampling and the publication of patient-level data so that interested researchers can replicate the results themselves or include them in future meta-analyses of the topic. The limitations of our article include the small number of treatment discontinuation events that prevented us from more sophisticated analyses or modeling. However, we instead validated existing prognostic models for the purpose of predicting treatment discontinuation.

## Conclusions

Treatment discontinuation of palliative brain radiotherapy in patients with brain metastases occurred in 7.5% of cases mostly due to clinical deterioration. Poor performance status as well as more advanced disease and, in turn, poor prognosis was associated with higher discontinuation rates.

## Data Availability

The full dataset, notebook and environment file have been uploaded to a public repository https://github.com/windisch-paul/rt-treatment-discontinuation.

